# Potential Impacts of Mass Nutritional Supplementation on Measles Dynamics: A Simulation Study

**DOI:** 10.1101/2021.09.10.21263402

**Authors:** Navideh Noori, Laura A. Skrip, Assaf P. Oron, Kevin A. McCarthy, Josh L. Proctor, Guillaume Chabot-Couture, Benjamin M. Althouse, Kevin P.Q. Phelan, Indi Trehan

## Abstract

The bidirectional interaction between undernutrition and infection can be devastating to child health. Nutritional deficiencies impair immunity and increase susceptibility to infection. Simultaneously, infections compound undernutrition by increasing metabolic demand, and impairing nutrient absorption. Treatment of acute malnutrition (wasting) can reverse some of its deleterious effects and reduce susceptibility to infectious diseases. Nutrition-specific approaches may be packaged with other interventions, including immunization, to support overall child health. To understand how mass nutritional supplementation, treatment of wasting, and vaccination affect the dynamics of a vaccine-preventable infection, we developed a population-level, compartmental model of measles transmission stratified by age and nutrition status. We simulated a range of scenarios to assess the potential reductions in measles infection and mortality associated with targeted therapeutic feeding for children who are wasted and with a mass supplementation intervention. Nutrition interventions were assumed to increase engagement with the health sector, leading to increased vaccination rates. We found that the combination of wasting treatment and mass supplementation coverage followed by an increase in vaccination coverage of non-wasted children from a baseline of 75% to 85%, leads to 34-57% and 65-77% reduction in measles infection and mortality and 56-60% reduction in overall mortality among wasted children, compared with the wasting treatment alone. Our work highlights the synergistic benefits that may be achieved by leveraging mass nutritional supplementation as a touch point with the health system, to increase rates of vaccination and improve child survival beyond what would be expected from the additive benefits of each intervention.

## Introduction

Undernutrition and infection interact in a bidirectional manner. Both micronutrient and macronutrient deficiencies stunt a child’s growth, impair immunity, increase susceptibility to infection, and worsen outcomes from common infectious diseases.^1^ The risk of adverse outcomes from infectionis correlated with the degree of undernutrition; concurrently, infection contributes to undernutrition by reducing a child’s ability to consume food, by contributing to nutrient malabsorption, and by increasing metabolic demand.^2-4^ As an example, a higher cumulative burden of diarrhea during the first two years of life increases the risk of stunting at 24 months of age. ^5^ Improving the baseline nutritional status of children and providing treatment for acute malnutrition (wasting) lowers the negative impacts of infections on growth by strengthening children’s immune systems, preventing poor appetite, compensating for malabsorption, and favoring the growth of beneficial gut microorganisms.^2,6^

Undernutrition can lead to long-lasting immune deficits, rendering undernourished children at a significantly increased risk of respiratory infections, diarrhea, malaria, and measles.^3, 7-9^ Even seemingly mild manifestations of these common infectious diseases can have long-term effects on children’s physical and cognitive development.^10^ Identifying patterns of association between undernutrition and infection are important to the clinical and public health efforts in reducing childhood morbidity and mortality.^1^ The bidirectional relationship between the two events has been well-documented, and its importance recognized for decades.^4,11^ Few studies, though, have explored the impact of undernutrition on the transmission dynamics of infectious diseases. Among these, there has been investigation into the effect of undernutrition, or specific nutritional therapies, on transmission of tuberculosis^12^ and cholera,^13^ emphasizing the need to address nutrition to reduce the burden of the infection. There are also animal models of infection and undernutrition that show infection can cause undernutrition; specifically, weanling undernutrition exacerbates infection and mucosal disruption,^14-17^ and increases the intensity of infection in neonatal mice, as assessed by stool shedding, by 1-4 orders of magnitude.^15^

Undernutrition is a primary contributor to death in 44.8% of childhood fatalities from measles.^18^ Malnourished children are more likely to develop complications of measles and have a higher case-fatality ratio,^19^ while measles infection can in turn worsen the nutritional status of children. Koster et al.^20^ found that measles had an adverse impact on both mortality and the nutritionalstatus of surviving children. In their prospective study in Bangladesh, children 7-23 months of age experienced a persistent nutritional deficit of about 10% after measles infection followed by prolonged diarrhea. Another study in an urban settlement in Guinea-Bissau also found a negative impact of measles on the nutritional status of children aged 9-35 months old.^21^ Several studies have shown that measles was a precipitating cause of kwashiorkor,^22-24^ and that there have been reports of a reduction in the incidence of kwashiorkor following measles vaccine campaigns.^23^

Interventions focused on treating or preventing undernutrition often rely upon increasing the engagement of mothers and children with the health sector. Intentionally packaging nutrition-specific care with routine immunization could lead to enhanced effectiveness in reducing undernutrition but also improve prevention efforts against infectious disease. In fact, nutritional commodities may be perceived as an incentive to draw patients to health centers where other services may be offered. Previous work has shown that an incentive as simple as a bag of lentils for each immunization visit, has large positive impacts on the uptake of immunization services in resource poor areas.^25^ In addition, vaccination uptake can enhance long-term nutrition outcomes, and targeted vaccination of children with poor socio-demographic characteristics can improve their overall nutrition status.^26^

We developed apopulation-level, dynamic model of measles transmission stratified by nutrition status to understand how nutrition-based interventions and vaccination collectively affect measles infection and mortality. In particular, we modeled two scenarios. In the first, wasted children received treatment with ready-to-use therapeutic food (RUTF). The second included treatment of wasted children, as well as mass supplementation of all children 6-23 months with small quantity lipid-based nutritional supplements (SQ-LNS). Lipid-based nutritional supplements (LNS) are a class of ready-to-use food supplements that are highly nutrient-dense and fortified with vitamins and minerals at levels designed to treat and prevent acute malnutrition. They range in ration sizes based on their use to either treat or prevent acute malnutrition, with ready-to-use therapeutic or supplementary foods (RUTF, RUSF) coming in 92- or 100-gram sachets, medium-quantity LNS in 50 gram sachets, and 20-gram sachets of SQ-LNS for home fortification of local diets to improve children’s complementary feeding. There is strong evidence in support of implementing mass SQ-LNS supplementation among at-risk children from 6-18 or 23 months, like those in Niger.^27^ Such supplementation has been shown to reduce stunting, wasting, anemia,^28^ and all-cause mortality^29^ as well as improve developmental outcomes.^30^

We accounted for the nature, timing, and magnitude of interactions between measles and undernutrition under the different simulated scenarios. Model parameters reflected the situation in Niger, which was considered to be an appropriate case study due to its high rates of undernutrition and its persistent measles burden. Undernutrition is the leading cause of infant mortality and morbidity in Niger.^31^ The levels of undernutrition in Niger are among the highest in the world, with 43.5% and 12.5% of children under 5 years of age being stunted and wasted in 2021, respectively; in the rural Zinder region, stunting and wasting rates were even higher at 57.4% and 14.3%.^32^ These rates are much higher than the average for the African continent as a whole (30.7% and 6.0% respectively).^33^ Outbreaks of vaccine-preventable diseases like measles are also common in Niger.^34,35^

Our modeling scenarios can help us understand the complex connections between undernutrition and measles that have not been disentangled by previous studies and may help inform approaches to intervention. There is strong evidence for the positive impact on preventing undernutrition and mortality through mass supplementation with child-adapted foods like SQ-LNS. If our model demonstrates an additive benefit of increased measles vaccination, it would justify further investment by governments and donors in scaling-up such programs especially in areas with a simultaneous high burden of malnutrition and low coverage for measles vaccination.

## Materials and Methods

### Measles-Undernutrition Model with Wasting Treatment (Scenario 1; Treatment)

We developed a seasonally-forced, deterministic, continuous-time Susceptible-Infectious-Recovered (SIR) model of measles transmission that accounts for vaccination and for nutritional interventions (Fig. 1). We stratified the population into children 6-23 months and individuals of all other ages, with children aging from the former into the latter compartment at a rate *α*. We assumed a fixed population of *N*_*0*_ = 100,000 and the demographic characteristics of Niger such as birth rate and death rates.^36-38^ The population size of infants up to 6 months is assumed to be *B* = 5000. A full list of parameter values is given in Table 1, and model equations are also given in supplementary material.

**Table 1.** Parameter values used in the dynamical measles-undernutrition model.

| Parameters | Symbol | Value | Reference |
| --- | --- | --- | --- |
| **Demographic and undernutrition parameters** |  |  |  |
| Population size | $N_0$ | 100,000 | - |
| Population of surviving infants under-6 months old | $B$ | 5,000 | - |
| Aging rate of age group 6-23 months | $\alpha$ | $1/(6+23)/2 \sim 1/15 \text{ month}^{-1}$ | - |
| Under-2 mortality rate | $\mu$ | 2.05 deaths per 10,000 per day $\sim 0.075$ per year | (36) |
| Proportion of infants under-6 months who are wasted | $\omega$ | 20% | (52) |
| Wasting mortality rate | $\mu_m$ | $0.016 + 0.075$ | (36, 53) |
| Wasting treatment coverage | $\tau$ | variable (defined as a rate) | (54) |
| Relapse rate | $\chi$ | 2% at 1 month post-discharge | (39) |
| **Measles parameters** |  |  |  |
| Measles basic reproductive ratio | $R_0$ | 15 | (55) |
| Seasonality amplitude of measles | $b_1$ | 0.6 | (45) |
| Force of infection | $\lambda$ | See Supplementary Material | - |
| Measles infectious period | $1/\gamma$ | 14 days | (45) |
| Immigration rate of measles | $\epsilon$ | 10 infected per year | (56) |
| Vaccination coverage of measles | $u$ | Defined as a rate to be equivalent to 75% | (40) |
| Vaccination coverage of wasted children | $uw$ | Defined as a rate to be equivalent to 67.5% | (41) |
| Vaccination coverage of children treated for wasting | $ut$ | variable (defined as a rate) | - |
| Measles case fatality rate | $\sigma$ | 2.8% | (57) |
| Increased measles mortality due to wasting | $\phi$ | $(6+3.7)/2 \sim 5$ | (7) |
| Increased susceptibility to measles infection due to wasting | $\theta$ | 2 | Assumed |
| Proportion of children who become wasted after measles infection | $H$ | 4% | (24, 42) |
| Increased susceptibility to measles due to subsequent wasting event | $\varsigma$ | 0 | Assumed |
| Measles-associated wasting mortality | $\rho$ | 44% | (42) |
| Infectious period extension for wasted children* | $\eta$ | Assuming infectious period among wasted children is 20 days, $\eta = 14/20 \sim 0.7$ | (58) |
| Proportion of wasted infected children who recover from wasting | $\delta$ | 20% | Assumed |
| **Mass nutritional supplementation parameters** |  |  |  |
| Mass supplementation coverage | $MC$ | 60% | Assumed |
| Measles vaccine coverage after mass supplementation | $u'$ | 75%, 80%, 85% | Assumed |
| Reduced all-cause mortality due to mass supplementation | $\mu m'$ | $0.075(1-0.27) + 0.016$ | (29) |
| Reduced probability of first AM event due to mass supplementation | $K$ | 29% | (43) |

\*Measles latent and infectious periods are approximately 6 and 8 days, respectively. Infectiousness starts about 4 days before the onset of rash and lasts ~ 4 days after the onset of rash.

**Figure 1.**
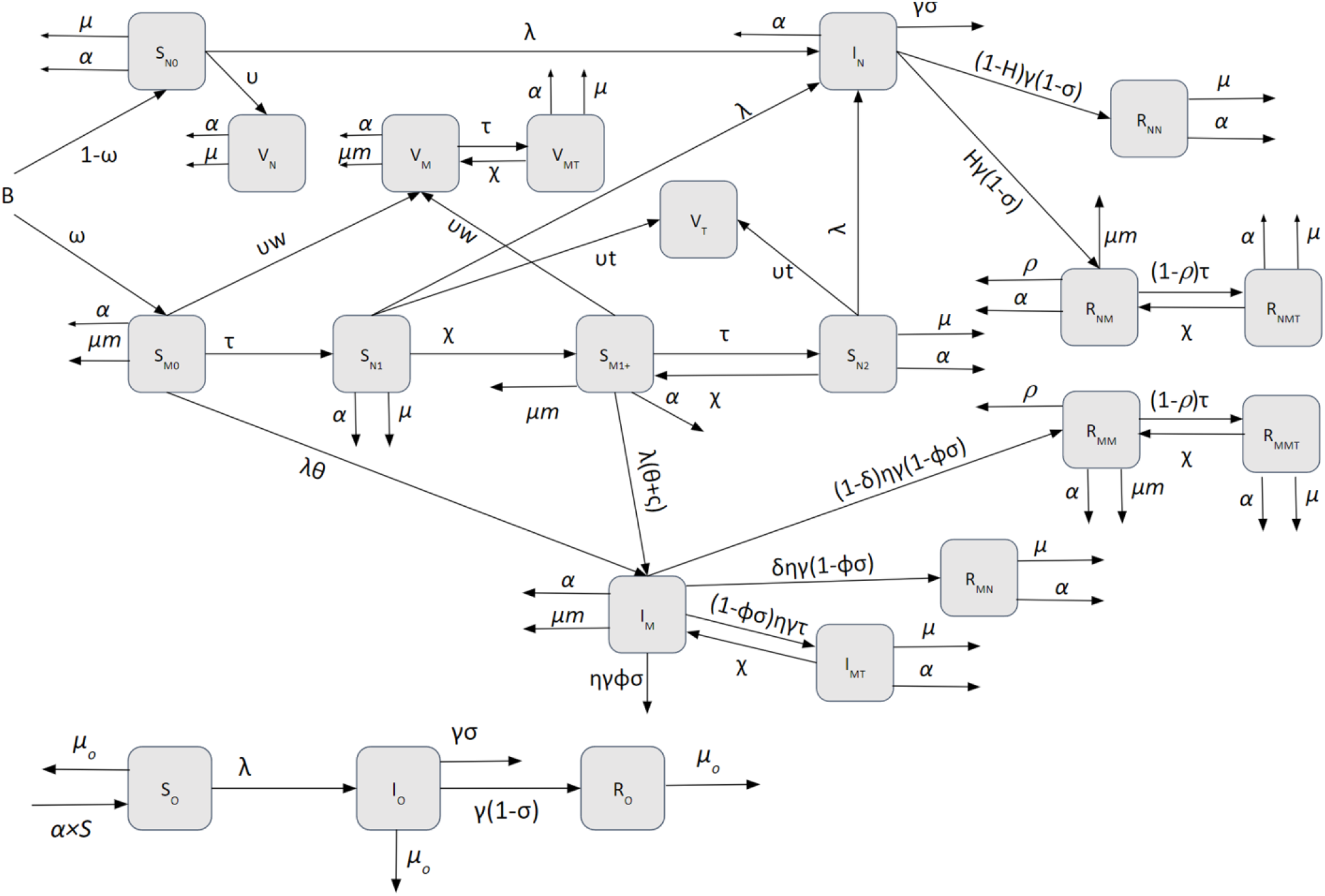
Schematic of measles-undernutrition model with no mass nutritional supplementation (Scenario 1).

Initially, children enter the two classes of susceptibility based on their nutritional status: nourished (non-wasted, *S*_*N0*_) or wasted (*S*_*M0*_). A proportion, *ω*, of children become wasted during the first 6 months of life. Some of the wasted children, regardless of their status in the model (susceptible to measles or vaccinated or recovered), receive therapeutic feeding and move to the non-wasted and treated compartments. The treatment rate of wasted children was varied to represent the coverage of 5% to 100%, with 75% effectiveness.. Previous studies have shown that after recovery from wasting, some children relapse and become wasted post-discharge.^39^ We accounted for relapse by moving treated individuals to different compartments at rate χ. The susceptible children who received treatment (S_N1_), were moved to compartment (S_M1+_). After individuals in *S*_*M1+*_ are treated again for wasting, they move to (*S*_*N2*_) and can in turn, move back to (*S*_*M1+*_) after experiencing relapse for the second time or more. We assume that wasted children have a higher mortality rate (*µm*) than in other states.

A proportion of individuals from all above-mentioned compartments are vaccinated and enter the vaccinated compartment (*V*). The vaccination coverage is assumed to be 75%, close to Niger national immunization coverage in 2016.^40^ We model single-dose vaccination and assume 95% of vaccinated children are no longer susceptible to measles infection. We also assumed that vaccination coverage of wasted children are 10% lower than nourished children (*υw* = 67.5%), based on the Demographic and Health Surveys data.^41^ The vaccination coverages in the model were defined as rates, and the proportion vaccinated was calculated from the model outputs. Based on observations of changes in overall care-seeking as a result of engagement of mothers through nutrition intervention programs,^25^ we hypothesize that receiving therapeutic feeding for wasting among children who are susceptible to measles, increases the measles vaccine uptake by *vt*.

Upon contracting measles, non-wasted and wasted individuals enter the infectious compartments (*I*_*N*_ and *I*_*M*_, respectively). Depending on the host age and condition, measles may be fatal. This is presented by the measles case fatality rate, here defined as probability (*σ*). We assume that wasting leads to increased susceptibility to measles (θ), longer infectious period (*η*), and increased measles mortality (ϕ). In this work, we assumed that the subsequent wasting event increases susceptibility to measles to the same degree as the first wasting event (*ς* = 0). Individuals who recover from measles move to the recovered class (*R*). A proportion, *H*, of children develop wasting after measles infection.^23^ Children who survive from measles but become wasted post-measles infection move from *I*_*N*_ to *R*_*NM*_, and those who stay nourished move to *R*_*NN*_ class. There is also a chance, *ρ*, of mortality due to measles-associated wasting.^24,42^ These parameter values were informed by literature on kwashiorkor specifically, due to a lack of studies on measles-associated marasmus. We assume that a proportion, *δ*, of wasted children who get infected with measles, eventually recover from wasting and move from *I*_*M*_ to *R*_*MN*_ upon infection. Those who stayed wasted, move from *I*_*M*_ to *R*_*MM*_. We assumed a proportion of individuals in *R*_*MM*_ experience death due to measles-associated wasting.

We depict the findings as heatmaps displaying measles infections averted across different combinations of therapeutic feeding rate for children with wasting (*τ*) and vaccination rate among wasted children receiving therapeutic feeding (*υt*). We vary these two quantities while fixing values of the remaining model parameters. The percentage reduction in infections and mortality due to measles among wasted children aged 6-23 months old, total infections and mortality in the population to measles among wasted children, and overall mortality among wasted children are the model outcomes. To calculate the percentage reduction, the sum of number of each outcome over the last year of simulations were calculated versus the baseline sum (*τ* = 0.05 and *υt* = 67.5). We also calculated the rates of infected cases and measles deaths per 100,000 wasted children and per 100,000 population per year, as well as rate of overall mortality per 100,000 wasted children.

### Measles-Undernutrition Model with Mass Nutritional Supplementation (Scenario 2; Prevention)

Instead of wasting treatment, we model the effect of community-wide mass nutritional supplementation on dynamics of measles by assuming that 60% of the population aged 6-23 months (parameter *MC*) receive daily doses of SQ-LNS (20 gr per sachet) at health center, per coverage found in a recent study in Mali^29^ (Table 3) (Fig. S1). We hypothesize that the mass supplementation results in an increase in the first dose of measles vaccine uptake in the region during the time the cohort receives supplementation. We represent this effect by conducting three scenarios and varying measles vaccination rate in a way that the estimated coverage changes from the baseline value of 75% to 80%, and 85% among susceptible individuals who are not wasted (*S*_*N0*_) after receiving the SQ-LNS. The increased measles vaccine uptake rate is shown as *υ’* (Fig. S1). Recent studies have shown significant improvement in children’s nutritional status as a result of receiving SQ-LNS. A cluster-randomized controlled trial in Mali has shown a 29% reduction in the probability of a first wasting event.^43^ We account for this effect in the model by parameter *k* (Fig. S1). Also, a recent pooled analysis of randomized controlled trials of LNS showed a 27% reduced probability of all-cause mortality.^29^ This effect is shown as *µm’* in the model.

The number of measles cases is calculated as the sum of (1) the number of measles cases where 40% of the population (*1-MC*) do not receive the mass supplementation, and (2) the number of measles case where 60% of population (*MC*) receive the mass supplementation, per unit time. The number of measles cases among wasted children, mortality due to measles among wasted children, and overall mortality among wasted children as well as their rates were calculated in a similar manner.

### Measles-Undernutrition Model with Mass Nutritional Supplementation and Wasting Treatment (Scenario 3; Treatment + Prevention)

We combine scenarios 1 and 2 and we model the combined effects of wasting treatment and the community-wide mass nutritional supplementation on dynamics of measles (Fig. S2).

### Sensitivity Analysis

To characterize the response of model outputs to parameter variations and to identify the influential parameters, we conducted a sensitivity analysis using Latin hypercube sampling-partial rank correlation coefficient (LHS-PRCC). LHS-PRCC is an efficient sampling-based sensitivity analysis that assumes the effect of a parameter on model outputs is monotonic after removing the linear effects of all parameters except the parameter of interest.^44^ Using LHS, we generated a matrix of 5000 sample points in the 9-dimensional unit cube (i.e., all parameters), and we explored how these parameters affected the model outcomes by using different simulated parameter sets. Next, we determined the effect of each parameter on the model outcomes while averaging over the other variables by inspecting the PRCC between each of the parameters and the percent change in different outcomes.

## Results

### Scenario 1

We modeled a 20% prevalence of wasting among children aged 6-23 months. Treating wasted children was assumed to increase vaccination coverage among wasted children who are susceptible to measles and seeking treatment. We first assumed a coverage of therapeutic treatment for wasted children to be 5% and explored the impact of incremental increases, along with constant or increased vaccination coverage among the treated children only, upon measles infection and measles mortality among wasted children and overall mortality. An underlying assumption is that unvaccinated wasted children who receive therapeutic feeding are given the measles vaccine during week 4 of treatment, as is indicated by most treatment protocols. Rates of treated and vaccinated individuals were converted to the proportions and shown in figures. Fig. 2 shows how the combination of wasting treatment and measles vaccination coverage impacts measles outcomes. Increasing the treatment coverage of wasted children from 5% to 95%, followed by vaccination coverage of children who received treatment from 5% to 95%, leads to up to a 36% reduction in measles cases (from 2673 to 2039 per 100,000 population), and 50% reduction in measles mortality (from 125 to 75 per 100,000 population) In addition to these benefits seen for the overall population of children, the combination of wasting treatment coverage from 5% to 95% followed by measles immunization coverage from 5% to 95% reduces the measles infection and mortality among wasted children specifically by 71%, and overall mortality of wasted children by 69% (Fig. S3).

**Figure 2.**
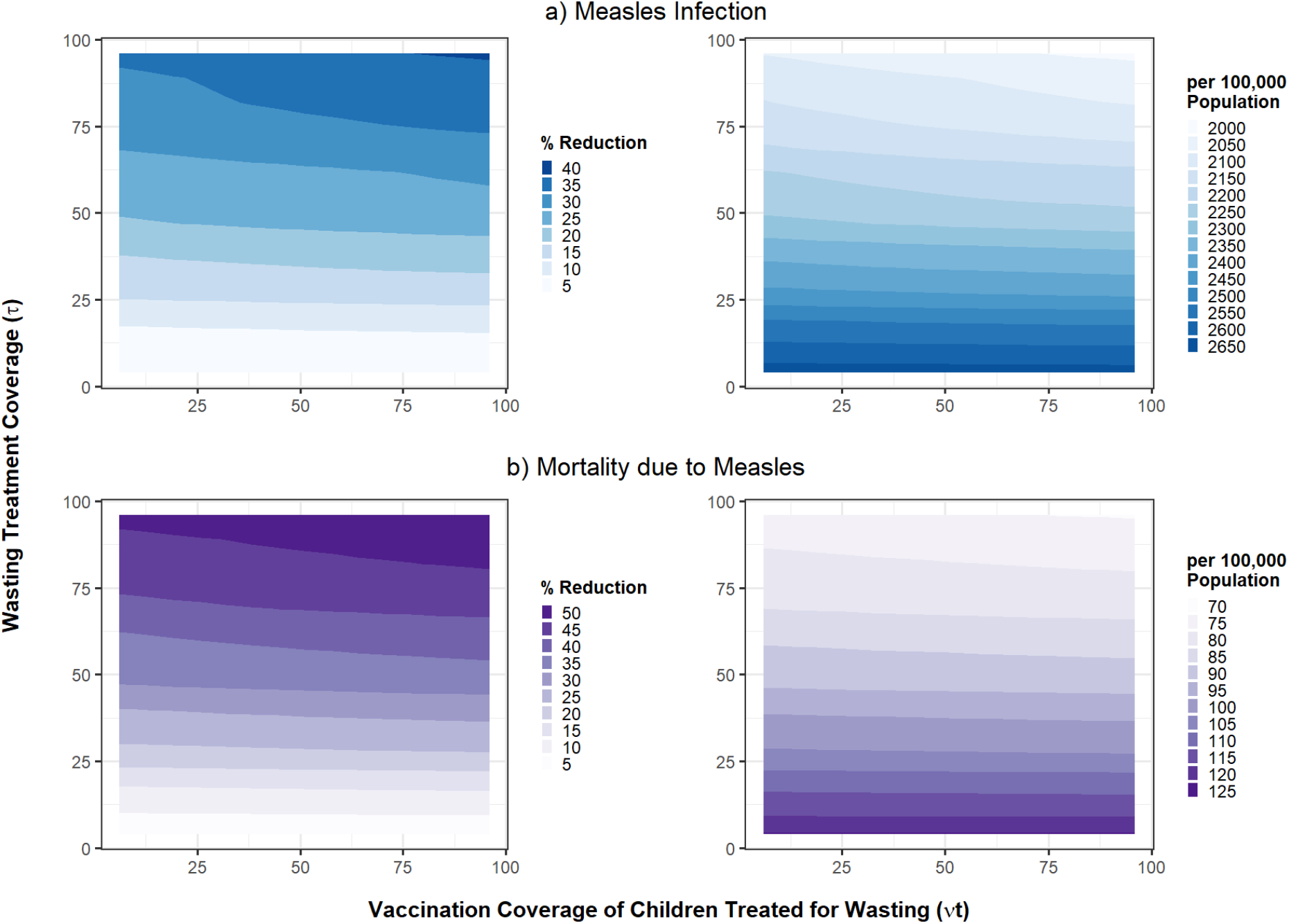
Combined impact of treatment of wasted children and their vaccination coverage (Scenario 1) on reduction in a) measles cases and b) mortality due to measles. The outcomes are shown as percentage reduction as well as incidence per 100,000 population.

### Scenario 2

Instead of therapeutic feeding of wasted children, we included mass supplementation for nutritionally at-risk populations - a less-targeted intervention with greater reach at the community-level. We assumed that 60% of the children 6-23 months old received mass supplementation with SQ-LNS at health centers and hypothesized that vaccination coverage of non-wasted children (*S*_*N*0_) who received supplementation increased from 75% to 85%. This led to a 20% reduction in measles infection and mortality in the population and 15% reduction among wasted children. It also leads to a 10% reduction in overall mortality among wasted children.

### Scenario 3

We combined Scenarios 1 and 2 and assessed the effects of therapeutic feeding of wasted children and mass supplementation of 60% of children 6-23 months old, together on measles outcomes. We varied the treatment coverage of wasted children (*τ*) between 5% to 95% as well as the vaccination coverage of treated children for wasting (*υt*) between 67.5% to 95%. An increase in vaccination coverage of non-wasted children as a result of mass supplementation (*υ’*) from 75% to 85%, leads to a 55% reduction in measles infection (from 1652 to 1246 per 100,000 population) and 66% reduction in measles mortality (from 42 to 24 per 100,000 population) (Fig. 3). The measles infection and mortality among wasted children aged 6-23 months old were reduced by 71% and overall mortality among wasted children was reduced by 67% (Fig. S4).

**Figure 3.**
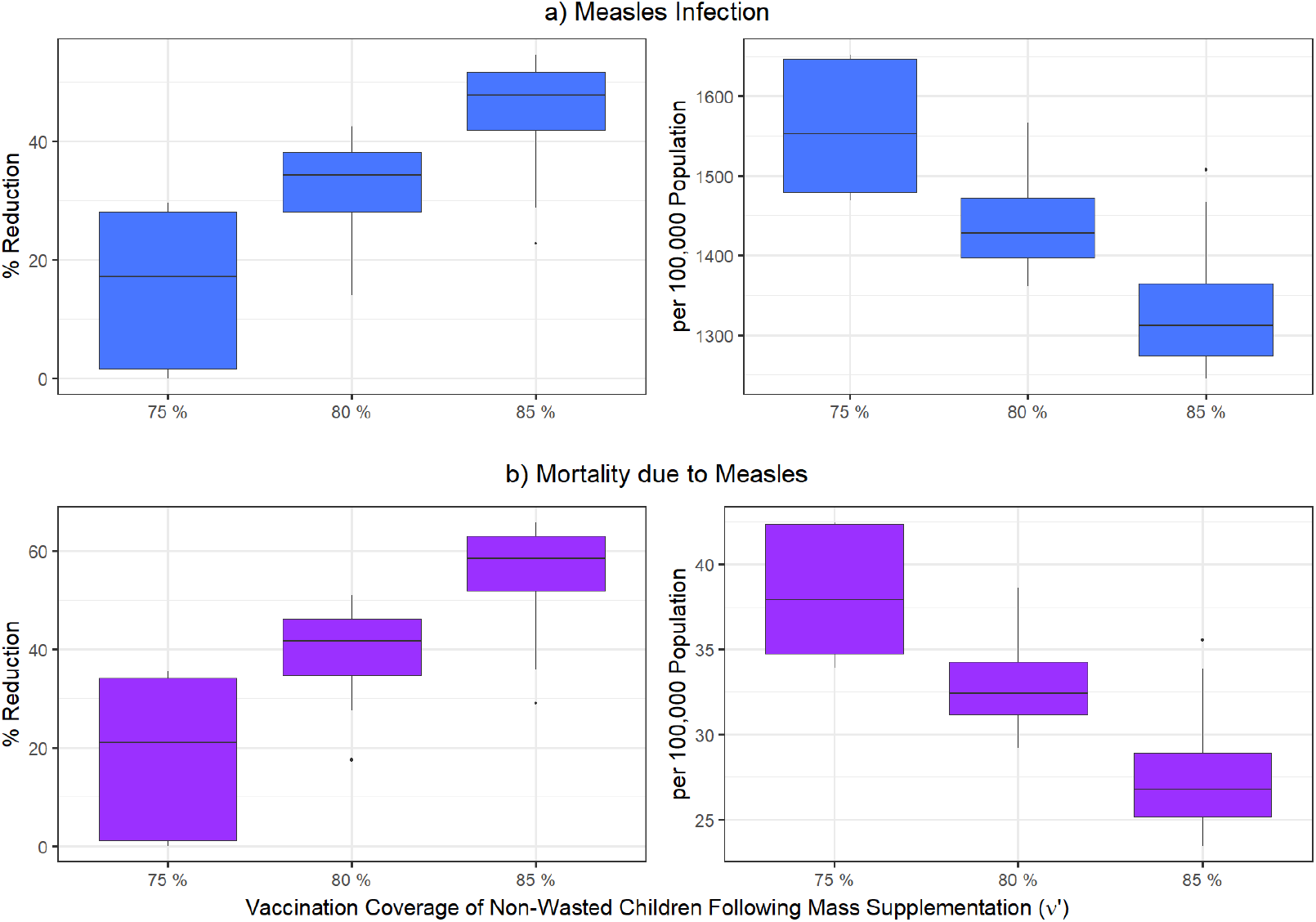
Impacts of mass nutritional supplementation and wasting treatment (scenario 3), on reducing a) measles infection (blue) and b) mortality due to measles (purple), assuming the vaccination coverage of non-wasted children (*υ’*) after receiving the SQ-LNS changes from its baseline value of 75% to 80%, and 85%. The variation in boxplots is produced by changing the treatment coverage of wasted children (*τ*) between 5% to 95%, and the vaccination coverage of wasted children between 67.5% to 95%.

Comparing Scenario 1 with Scenario 3 reveals that the absolute numbers of measles cases and measles mortality are lower in Scenario 3 than Scenario 1. 85% vaccination coverage of non-wasted children following mass supplementation, in addition to varying wasting treatment and vaccination coverage of children treated for wasting, leads to 36-57% and 67-77% reductions in measles infection and mortality, respectively, compared with wasting treatment only (Fig. 4 and Fig. S5). Among wasted children specifically, the infection and mortality due to measles also reduced by 74-78%, and overall mortality by 56-60%, for the 85% vaccination coverage (Fig. S6 and Fig.S7).

**Figure 4.**
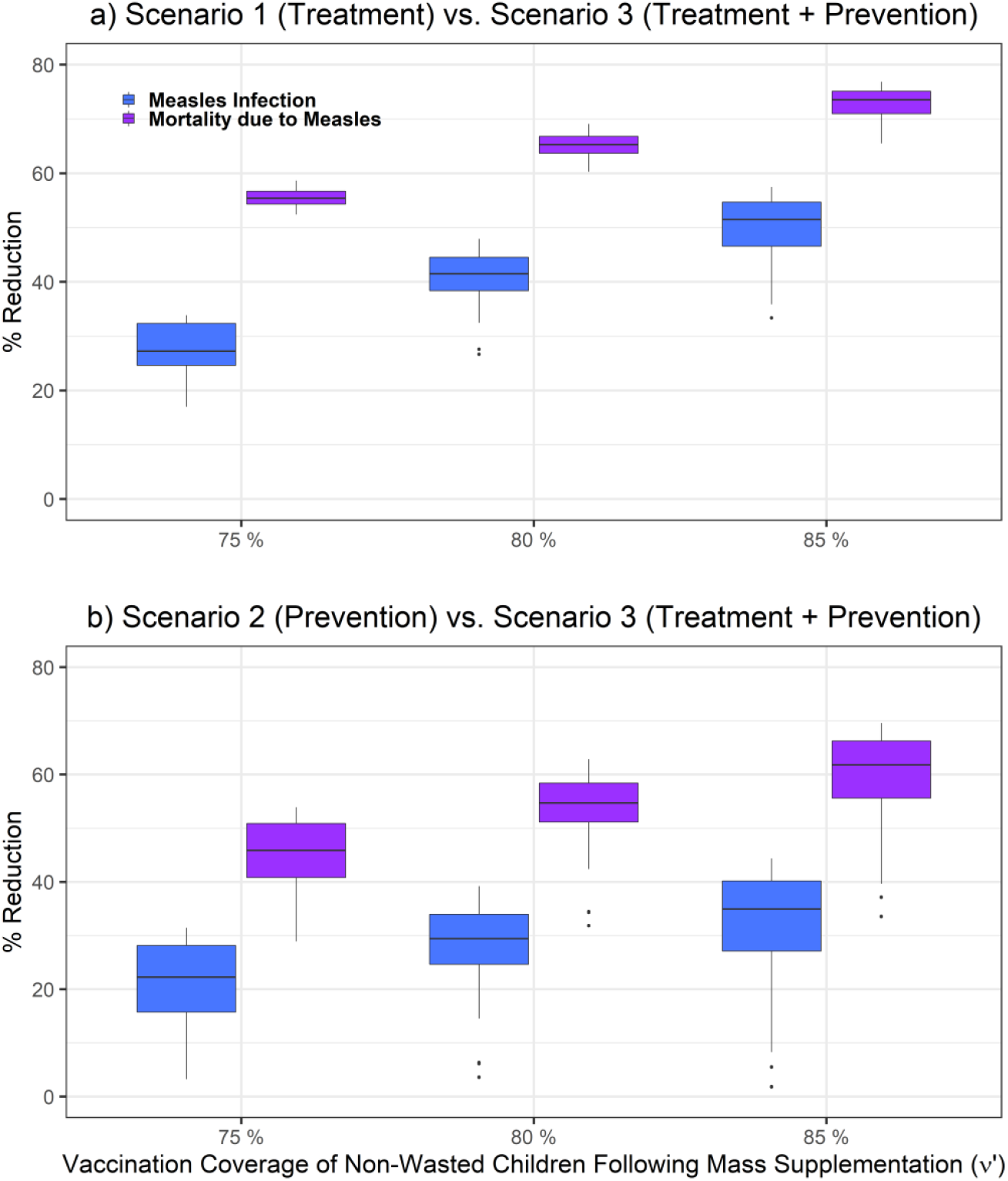
Difference between a) wasting treatment (Scenario 1) and mass nutritional supplementation and wasting treatment (Scenario 3) and b) mass nutritional supplementation (Scenario 2) and scenario 3, in reducing measles infection (blue) and mortality due to measles (purple), assuming the vaccination coverage of non-wasted children (*υ’*) after receiving SQ-LNS varies to 75%, 80%, and 85%. The variation in boxplots is produced by changing the treatment coverage of wasted children (*τ*) between 5% to 95%, and the vaccination coverage of wasted children between 67.5% to 95%.

If mass supplementation leads to 85% vaccination coverage of non-wasted children in both Scenarios 2 and 3, comparing Scenario 3 outcomes with Scenario 2 shows 5-44% reduction in measles infection and 37-70% reduction in measles mortality (Fig. 4 and Fig S.5). The variation in boxplots is produced by changing the treatment coverage of wasted children as well as vaccination coverage of children who received treatment for wasting. Increasing the = treatment coverage of wasted children increases the effect size of Scenario 3 in comparison to Scenario 2 and leads to larger reduction in measles infection, measles mortality and overall mortality (Fig. S6 and Fig. S7).

Overall, comparing the three scenarios shows that prevention could be a more effective approach than treatment. Assuming that in the prevention scenario 75% of non-wasted children who received nutrition supplementation received vaccination, and in the treatment scenario, 75% of wasted children receive treatment, and 75% vaccination coverage of treated children, number of infected cases per 100,000 wasted children reduced from 13574 (Scenario 1. treatment scenario) to 6260 (Scenario 2. prevention scenario) and to 4929 (Scenario 3. treatment + prevention scenario). Number of measles deaths per 100,000 wasted children was also reduced from 1900 (treatment scenario) to 876 (prevention scenario) and to 690 (treatment + prevention scenario) (Fig. S9). Similar results were obtained for the measles infection as well as measles mortality per 100,000 population (Fig. S8). Provision of mass nutritional supplementation reduced the number of wasted children and therefore, the number of wasted infected children. This effect was much larger than the treatment effect. Scenarios 2 and 3 have similar impact sizes on measles burden, emphasizing that prevention is a more effective approach than treatment (Fig. S8 and Fig. S9).

These differences in model outcomes were estimated by fixing the input parameter values to best estimates informed by the literature and to represent the dynamics of measles in the Zinder region of Niger^45^ (Table 1). However, precise quantities to inform some of the model parameters are unknown or unavailable, specifically, the increase in infectious period and increase in susceptibility to measles due to wasting (*η*, θ). Simulation over a range of values for one parameter with others fixed allows us to examine the effect of Scenario 1 on multiple model outcomes. As the magnitude of wasting effects – such as susceptibility effect and duration of infectious period – get larger, they result in an increase in the number of infected children, and number of deaths per 100,000 wasted children aged 6-23 months old, assuming the vaccination coverage of children who received treatment varied between 67.5% to 100% and the wasting treatment coverages varies between 5% to 100% (Fig. S10 and Fig. S11). For instance, an increase in susceptibility effect from θ = 2 to θ = 4, leads to, on average, a 50% increases in measles infection among wasted children (from 13988 to 22085 per 100,000 wasted children aged 6-23 months old per year) (Fig. S10). Varying other parameter values such as increased measles mortality due to wasting (ϕ), or proportion who recover from wasting after measles infection (δ) does not lead to a noticeable difference in the simulation results for the number of infected cases. Further sensitivity analyses using the LHS-partial rank correlation coefficient (LHS-PRCC) index and generating a matrix of 5000 sample points in the 9-dimensional unit cube (all parameters) indicate that *σ* (measles-induced mortality probability) θ (susceptibility impact of measles infection) have the strongest influence on both measles infection and measles mortality outcomes (Fig. S12). In addition, mortality due to measles among wasted children aged 6-23 months old is sensitive to ϕ (increased measles mortality due to wasting).

## Discussion

Substantive barriers to improving child health in resource-limited settings include persistently high rates of undernutrition and low rates of vaccine coverage for common infections such as measles. Given the close bidirectional relationship between childhood undernutrition and infectious diseases among vulnerable populations, there may be opportunities for synergy wherein the treatment and/or prevention of one condition leads to decreased rates of the other. Our work aims to capture the dynamic interactions between undernutrition and measles, so as to estimate the impact of treatment of wasting, mass nutritional supplementation for nutritionally at-risk populations, and measles vaccine coverage on measles infection and mortality.

No other nutrition intervention has shown such significant, simultaneous impacts on childhood malnutrition as mass supplementation with SQ-LNS from 6-18 or -23 months, including reductions in wasting, stunting, anemia, and mortality as well as improved developmental outcomes.^27,28,29,30^ Our models demonstrate a strong effect of mass nutritional supplementation in reducing measles infection and death, as well as overall mortality, if such an intervention could be leveraged to incentivize measles vaccination uptake. Importantly, modestly increasing measles vaccination coverage by providing vaccination at the same time and location as blanket nutritional supplementation leads to sizeable reductions in measles cases and mortality well above treatment of wasting or increased vaccine uptake alone. In addition to the synergic effect, our dynamical model demonstrates the nonlinear impacts of these interventions on measles burden.

Our results show that mass supplementation alone and mass supplementation combined with wasting treatment have a similar effect size on reducing measles infection and mortality. Mass supplementation with SQ-LNS coupled with vaccination would likely be more cost-effective than the individual treatment of wasting alone which, in high burden countries, puts financial pressures on vulnerable and poorly resourced health systems. Future work should explore the cost-effectiveness of integrated interventions. We used model parameters based on data from Niger, a nation plagued by a high burden of undernutrition, where interventions focused on the early childhood nutrition have the potential for a large impact on measles infection rates. Niger has made very little progress towards achieving international targets for decreasing stunting and wasting.^46^ Our results suggest that mass nutritional supplementation among vulnerable populations using SQ-LNS has over twice the effect on reducing measles infection and mortality as well as overall mortality among wasted children, compared with treatment of wasted children alone, assuming both methods lead to increased vaccination coverage. Therefore, expanding the reach of mass nutritional supplementation, including providing nutrition supplementation during routine immunization visits, could help Niger make progress towards lowering the incidence of wasting, as well as indirectly improve vaccination coverage. The models could be applied to other countries like Chad, Nigeria, or the Democratic Republic of Congo which have a similar burden of undernutrition as Niger but much lower measles vaccination coverage.

As with all models, there are some limitations to our work. Our work is theoretical, however our sensitivity analyses show that our model is robust to choice of parameter values, and thus is a call for more empirical studies. Although we account for multiple interactions between measles and undernutrition, there are several features of the complex system of measles epidemiology that were not included in the model. For instance, we did not account for any potential impact of undernutrition on measles vaccine leading to a potential delay in response to vaccine or waning of vaccine, as it is under-explored in the literature.^47^ In addition, one of the complications that malnourished children experience following measles infection, is post-measles diarrhea which exacerbates nutrient loss.^1^ Accounting for this could help us better identify the impacts of infection on children’s nutritional status. Also, among surviving children following measles infection a certain percentage experience blindness, a certain percentage with some residual pneumonia, and quite significantly a certain percentage with a residual post-measles immunodeficiency. The reported deaths due to measles do not account for mortality from these various comorbidities.^48-49^ In addition, assessing the potential increased pathogenicity of measles as a result of undernutrition would be useful in understanding the cyclic relationship between the two events to develop more effective interventions. More field data to constrain model parameters around the relationship between measles infection and undernutrition would allow for increasingly rigorous quantification of direct and indirect impacts of nutrition interventions. Also, nutrition interventions that led to rises in vaccination coverage were assumed to be delivered at health centers and at regular intervals. It will also be important to investigate different distribution models for supplementation in order to retain contact with health personnel without placing undue burden on already overworked caretakers.

Given the complex interactions between undernutrition and infectious diseases, it is still challenging to recognize where and at what level specific interventions can be most effective. The large contribution of undernutrition and infection to the global burden of disease increases the importance of studying these two processes. Our study offers a more nuanced understanding of the complex interactions between measles transmission dynamics and undernutrition and helps motivate advocacy for nutrition-focused interventions based on their multidimensional impacts. This work also helps understand how the addition of nutritional status, preventive nutrition interventions, and therapeutic treatments into mathematical models of vaccine-preventable diseases can better explain increased susceptibility to infection despite achieving certain vaccine coverage levels. This model also has implications for understanding other indirect benefits of increased care-seeking due to engagement with the health system for nutrition-based interventions. When considering how health systems with limited budgets should spend their money to achieve maximal benefit, we have shown here that leveraging mass nutritional supplementation as a contact point with the health system to even modestly increase measles vaccination coverage has a synergistic benefit beyond either intervention alone. Such a double impact could be even more important in areas where both malnutrition and measles are rising because of conflict, climate shocks, and disruptions from the Covid-19 pandemic.^50,51^

## Data Availability

There are no data underlying this work.

## Acknowledgments

The funders had no role in study design, data collection, data analysis, the decision to publish, or preparation of the manuscript. The content of this article is solely the responsibility of the authors and does not necessarily represent the official views of their respective employers or funders.

## Authors’ Contributions

IT, KPQP contributed to the concept, design and interpretation of the analysis. NN and LS and JLP developed the model. NN conducted the analysis. KAM, JLP, GCC, and BMA contributed to the model revision. NN, LS, APO, KAM, JLP, GCC, BMA, IT, KPQP wrote the manuscript.

