## Supplementary Material for "Potential Impacts of Mass Nutritional Supplementation on Measles Dynamics: A Simulation Study"

### Supplementary Information for
### Potential Impacts of Mass Nutritional Supplementation on Dynamics of Measles: A Simulation Study


Navideh Noori[a,*], Laura Skrip[b], Assaf P. Oron[c], Kevin A McCarthy[a], Joshua L Proctor[a], Guillaume Chabot-Couture[a], Benjamin M. Althouse[d,e], Kevin P.Q. Phelan[f,§], and Indi Trehan[g,§]

[a]Institute for Disease Modeling, Global Health Division, Bill and Melinda Gates Foundation, 500 5th Ave N, Seattle, WA, 98109, USA

[b]School of Public Health, University of Liberia, 1000 Monrovia, 10 Liberia

[c]Institute for Health Metrics and Evaluation, University of Washington, Seattle, WA, USA

[d]Information School, University of Washington, Seattle, WA, 98104, USA

[e]Department of Biology, New Mexico State University, Las Cruces, NM, 88003, USA

[f]The Alliance for International Medical Action (ALIMA), Paris, 75011, France

[g]Department of Pediatrics, Global Health, and Epidemiology, University of Washington, Seattle, WA, 98105, USA

[§]Kevin P.Q. Phelan and Indi Trehan are joint senior authors.


**This PDF file includes:**

Supplementary text

Table S1

Figures S1 to S12

### Supplementary Information Text

#### Measles-Undernutrition Model with Wasting Treatment (Scenario 1)

#### Age group 6-23 months old

$$\frac{dS_{N0}}{dt} = 2 \times (1-\omega)B - \lambda S_{N0} - 0.95 \times \nu S_{N0} - (\alpha + \mu)S_{N0} \tag{1}$$

$$\frac{dS_{M0}}{dt} = 2 \times \omega B - \theta\lambda S_{M0} - 0.95 \times \nu w S_{M0} - 0.75 \times \tau S_{M0} - (\alpha + \mu m)S_{M0} \tag{2}$$

$$\frac{dS_{N1}}{dt} = 0.75 \times \tau S_{M0} - \lambda S_{N1} - 0.95 \times \nu t S_{N1} - (\chi + \alpha + \mu)S_{N1} \tag{3}$$

$$\frac{dS_{M1+}}{dt} = \chi(S_{N1} + S_{N2}) - (\theta + \varsigma)\lambda S_{M1+} - 0.95 \times \nu w S_{M1+} - 0.75 \times \tau S_{M1+} - (\alpha + \mu m)S_{M1+} \tag{4}$$

$$\frac{dS_{N2}}{dt} = 0.75 \times \tau S_{M1+} - \lambda S_{N2} - 0.95 \times \nu t S_{N2} - (\chi + \alpha + \mu)S_{N2} \tag{5}$$

Non-wasted who received vaccinated

$$\frac{dV_N}{dt} = 0.95 \times \nu S_{N0} - (\alpha + \mu)V_N \tag{6}$$

Wasted who received treatment and then vaccinated

$$\frac{dV_T}{dt} = 0.95 \times \nu t(S_{N1} + S_{N2}) - (\alpha + \mu)V_T \tag{7}$$

Wasted who received vaccinated

$$\frac{dV_M}{dt} = 0.95 \times \nu w(S_{M0} + S_{M1+}) + \chi V_{MT} - 0.75 \times \tau V_M - (\alpha + \mu)V_M \tag{8}$$

Wasted who received vaccinated then received treatment

$$\frac{dV_{MT}}{dt} = 0.75 \times \tau V_M - (\chi + \alpha + \mu)V_{MT} \tag{9}$$

$$\frac{dI_N}{dt} = \lambda(S_{N0} + S_{N1} + S_{N2}) - (\gamma + \alpha + \mu)I_N \tag{10}$$

$$\frac{dI_M}{dt} = \theta\lambda S_{M0} + (\theta + \varsigma)\lambda S_{M1+} - (\eta \times \gamma + \alpha + \mu m)I_M + \chi I_{MT} - (1 - \phi \times \sigma)\eta \times \gamma \times 0.75 \times \tau I_M \tag{11}$$

$$\frac{dI_{MT}}{dt} = (1 - \phi \times \sigma)\eta \times \gamma \times 0.75 \times \tau I_M - (\chi + \alpha + mu)I_{MT} \tag{12}$$

$$\frac{dR_{NM}}{dt} = H(1-\sigma)\gamma I_N + \chi R_{NMT} - 0.75 \times \tau(1-\rho)R_{NM} - (\rho + \alpha + \mu m)R_{NM} \tag{13}$$

$$\frac{dR_{NMT}}{dt} = 0.75 \times \tau(1-\rho)R_{NM} - (\chi + \alpha + \mu)R_{NMT} \tag{14}$$

$$\frac{dR_{NN}}{dt} = (1-H)(1-\sigma)\gamma I_N - (\alpha + \mu)R_{NN} \tag{15}$$

$$\frac{dR_{MN}}{dt} = \delta(1-\phi \times \sigma)\eta\gamma I_M - (\alpha + \mu m)R_{MM} \tag{16}$$

$$\frac{dR_{MM}}{dt} = (1-\delta)(1-\phi \times \sigma)\eta\gamma I_M + \chi R_{MMT} - 0.75 \times \tau(1-\rho)R_{MM} - (\rho + \alpha + \mu m)R_{MM} \tag{17}$$

$$\frac{dR_{MMT}}{dt} = 0.75 \times \tau(1-\rho)R_{MM} - (\chi + \alpha + \mu)R_{MMT} \tag{18}$$

#### Age group 24 months and older

$$\frac{dS_o}{dt} = \alpha(S_{N0} + S_{M0} + S_{M1+} + S_{N1} + S_{N2}) - \lambda S_o - \mu o S_o \tag{19}$$

$$\frac{dI_o}{dt} = \lambda S_o - \gamma I_o - \mu o I_o \tag{20}$$

$$\frac{dR_o}{dt} = (1-\sigma)\gamma I_o - \mu o R_o \tag{21}$$

We developed a seasonally forced deterministic continuous-time SIR model. The seasonally forced system in our model is analyzed by making the transmission rate vary cosinusoidal which is given in the equations 22 and 23. $\lambda$ is the force of infection. $\epsilon$ is number of imported cases from outside the population.The mean transmission rate is given by the parameter $\beta$, with the amplitude of seasonality $b_1$. $\beta_0$ is the mean transmission rate which is obtained based on the $R_0$ equation. $\mu$, $\mu m$ and $\mu o$ denote constant *per capita* death rates, and $\omega$ represents proportion of infants under-6 months with overall wasting.

$$\lambda(t) = \beta(t)(I_N + I_M + I_o + \epsilon)/N \tag{22}$$

$$\beta(t) = \beta_0(1 + b_1 cos(2\pi t)) \tag{23}$$

$$R_0 = \frac{\beta_0}{\omega\eta\gamma + (1-\omega)\gamma + \mu + \mu m + \mu o} \tag{24}$$

#### Measles-Undernutrition Model with Mass Nutritional Supplementation (Scenario 2)

We remove the wasting treatment and instead, model the effect of mass nutritional supplementation on the dynamics of measles by assuming that 60% of the population aged 6-23 months (shown as parameter $MC$) receive SQ-LNS (Fig. S1). We assume the rest of population $(1 - MC)$ do not receive the mass supplementation. The $R_0$ equations for the $MC = 60$% of population with mass supplementation and rest of population (40%) are shown in equations 25 and 26, respectively:

$$R_0 = \frac{\beta_0}{MC(1-K)\omega\eta\gamma + MC(1-(1-K)\omega)\gamma + \mu + \mu m + \mu o} \tag{25}$$

$$R_0 = \frac{\beta_0}{(1-MC)\omega\eta\gamma + (1-MC)(1-\omega)\gamma + \mu + \mu m + \mu o} \tag{26}$$

##### Measles-Undernutrition Model with Mass Nutritional Supplementation and Wasting Treatment (Scenario 3)

We combine scenarios 1 and 2, and model the effect of mass nutritional supplementation as well as wasting treatment on the dynamics of measles by assuming that 60% of the population aged 6-23 months receive SQ-LNS (Fig. S2). All the equations are same as the previous model, only the following ones change and also $\mu m$ and $\nu$ change to $\mu m'$ and $\nu'$, respectively.

$$\frac{dS_{N0}}{dt} = 2 \times MC(1-(1-K)\omega)B - \lambda S_{N0} - 0.95 \times \nu' S_{N0} - (\alpha+\mu)S_{N0} \tag{27}$$

$$\frac{dS_{M0}}{dt} = 2 \times MC(1-K)\omega B - \theta\lambda S_{M0} - 0.95 \times \nu w S_{M0} - 0.75 \times \tau S_{M0} + (\alpha+\mu m')S_{M0} \tag{28}$$

**Table S 1.** Range of parameter values for the sensitivity analysis using scenario 1.

| Parameters | Range |
|---|---|
| Relapse rate ($\chi$)* | c(0.1,5) |
| Infectious period extension for wasted children $\eta$) | c(0.01,1) |
| Increased susceptibility to measles infection due to wasting ($\theta$) | c(1,10) |
| Measles case fatality rate ($\sigma$) | c(0.01,0.2) |
| Increased susceptibility to measles due to subsequent wasting event ($\varsigma$) | c(0,10) |
| Measles-associated wasting mortality ($\rho$) | c(0.01,1) |
| Proportion of children who become wasted after measles infection (H) | c(0.01,0.9) |
| Increased measles mortality due to wasting ($\phi$) | c(1,5) |
| Proportion of wasted infected children who recover from wasting ($\delta$) | c(0.01,0.5) |

* In annual scale

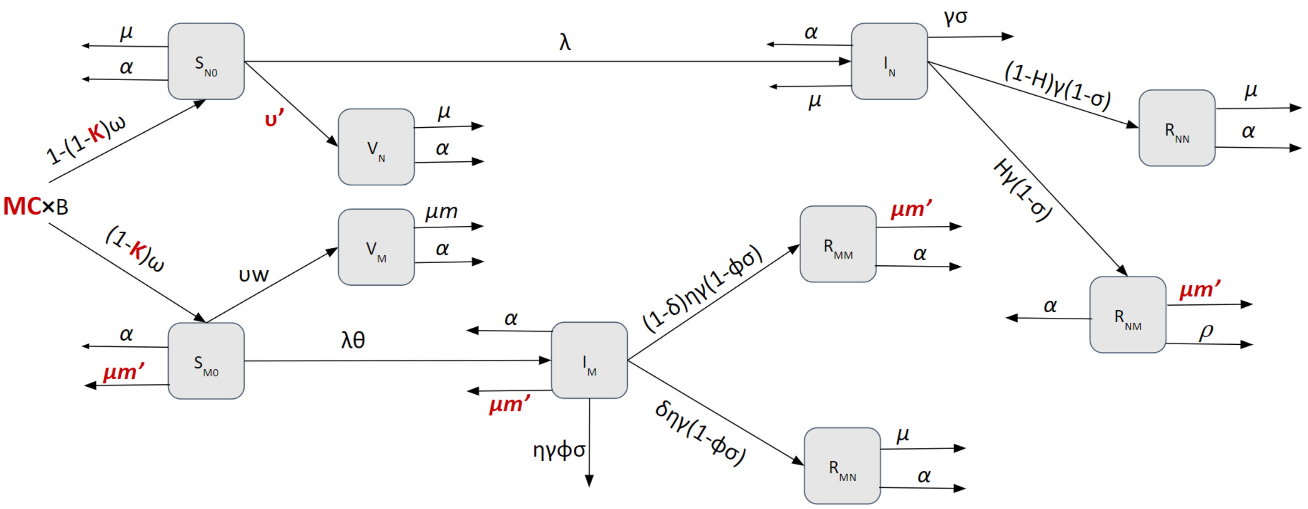


**Figure S 1.** Schematic of measles-undernutrition model with mass nutritional supplementation (Scenario 2).

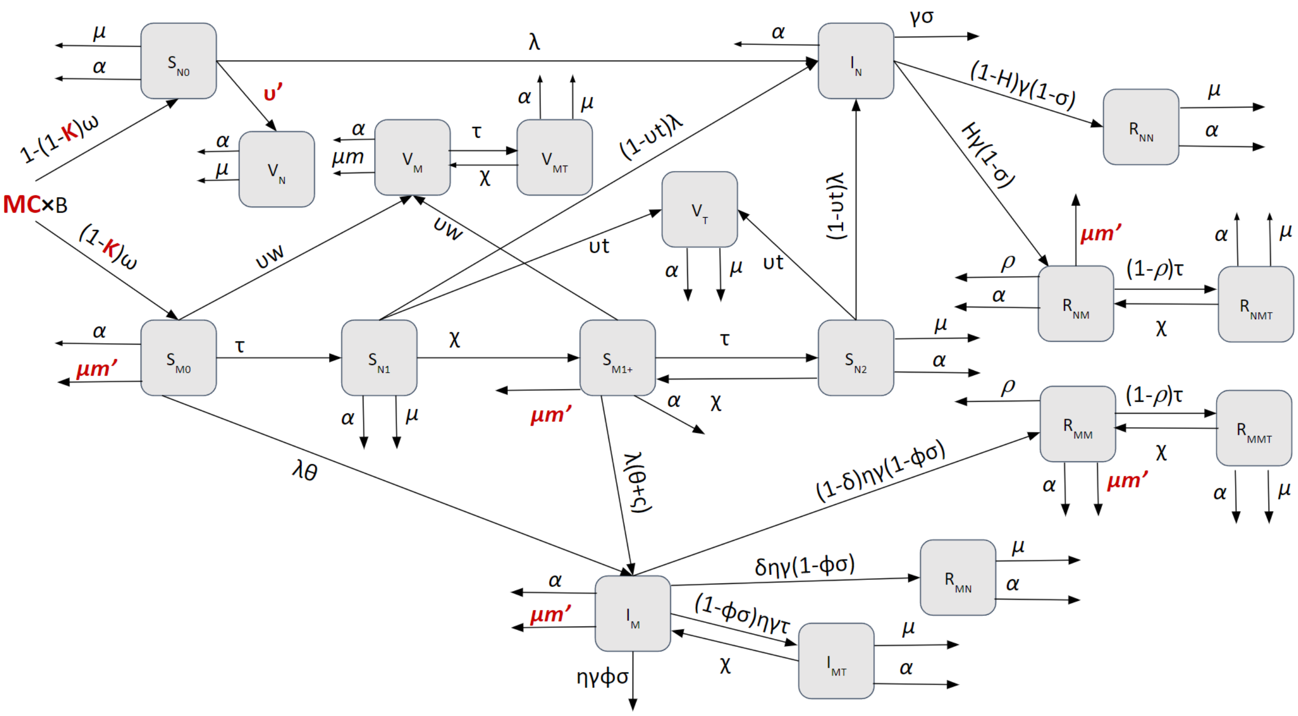


**Figure S 2.** Schematic of measles-undernutrition model with wasting treatment and mass nutritional supplementation (Scenario 3).

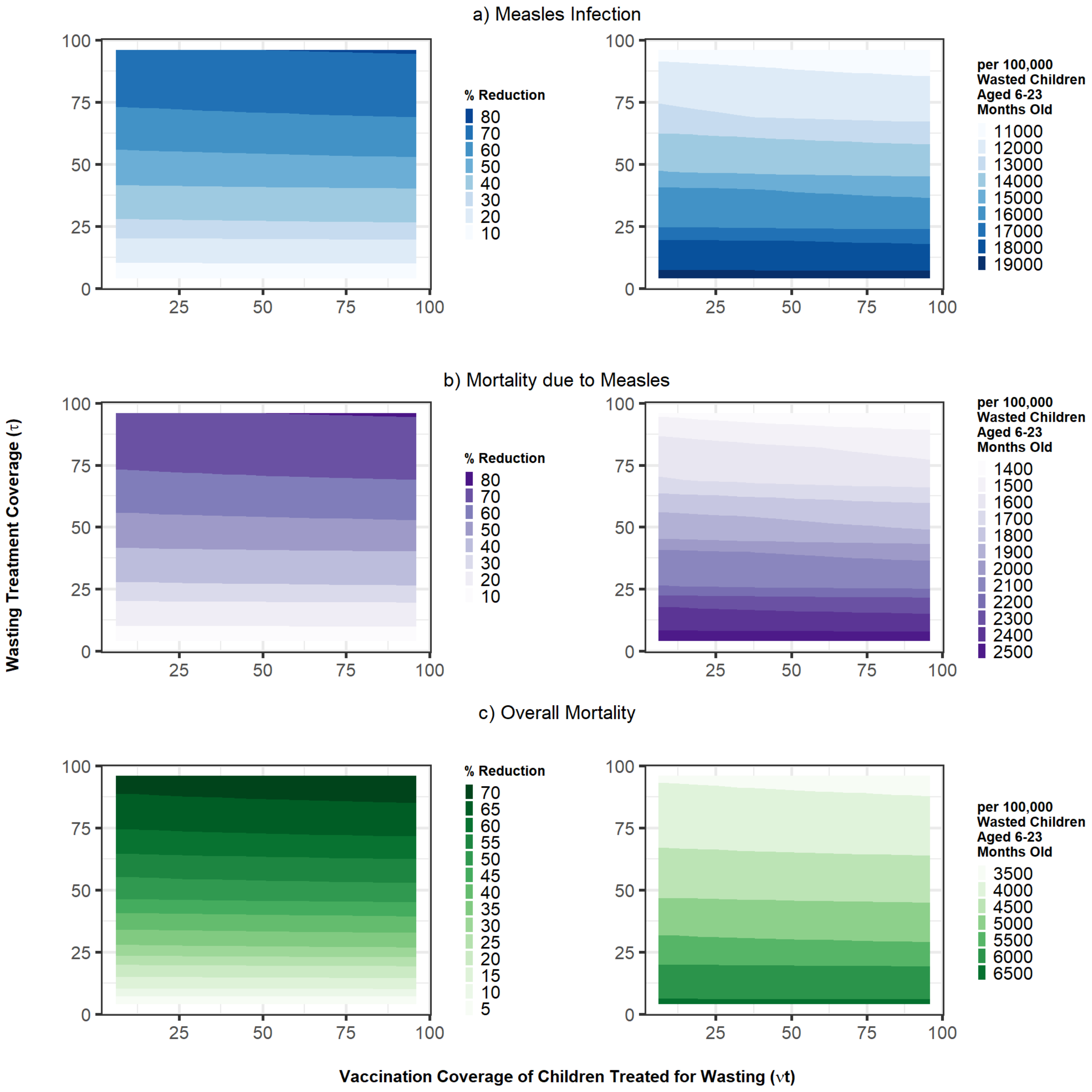


**Figure S 3.** Impact of wasting treatment and vaccination coverage of wasted children (scenario 1) on reducing a) measles infection and b) mortality due to measles among wasted children, and c) overall mortality among wasted children. The impact size is shown as percentage reduction in each outcome as well as incidence per 100,000 wasted children aged 6-23 months old.

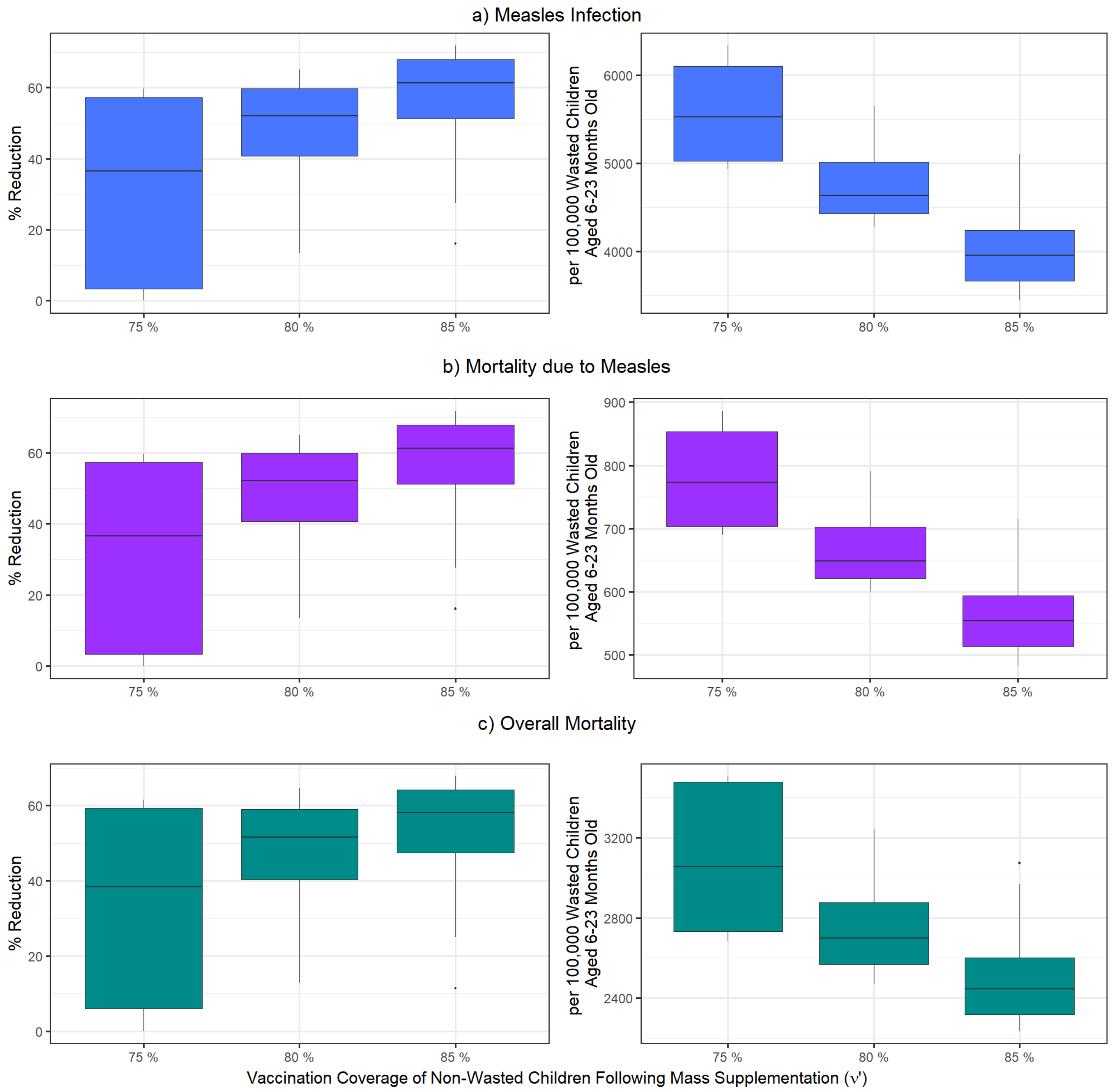


**Figure S 4.** Impact of mass nutritional supplementation + wasting treatment (scenario 3) on reducing measles infection and mortality due to measles among wasted children aged 6-23 months old, and overall mortality among wasted children. The impact size is shown as percentage reduction in each outcome as well as incidence per 100,000 wasted children aged 6-23 months old. The variation in boxplots is produced by changing the treatment coverage of wasted children and vaccination coverage of children who received treatment for wasting.

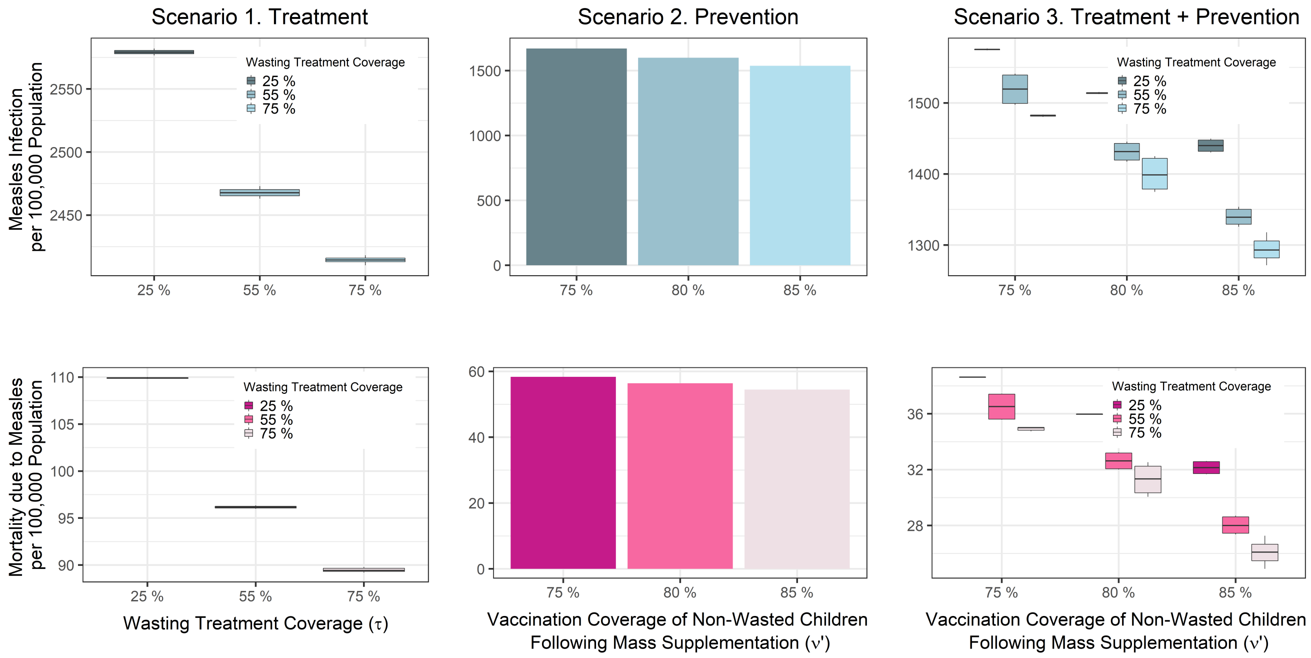


**Figure S 5.** Comparison of measles infection per 100,000 population and mortality due to measles per 100,000 population across three scenarios. Scenario 1 shows changes in outcomes by wasting treatment coverage of ($\tau$: 25%, 55% and 75%), and scenario 2 shows changes in outcomes by vaccination coverage of non-wasted children following mass supplementation ($\nu'$: 75%, 80%, 85%). Scenario 3 shows changes in outcomes by varying both $\tau$ and $\nu'$. The variation in boxplots is produced by changing the vaccination coverage of children who received treatment for wasting.

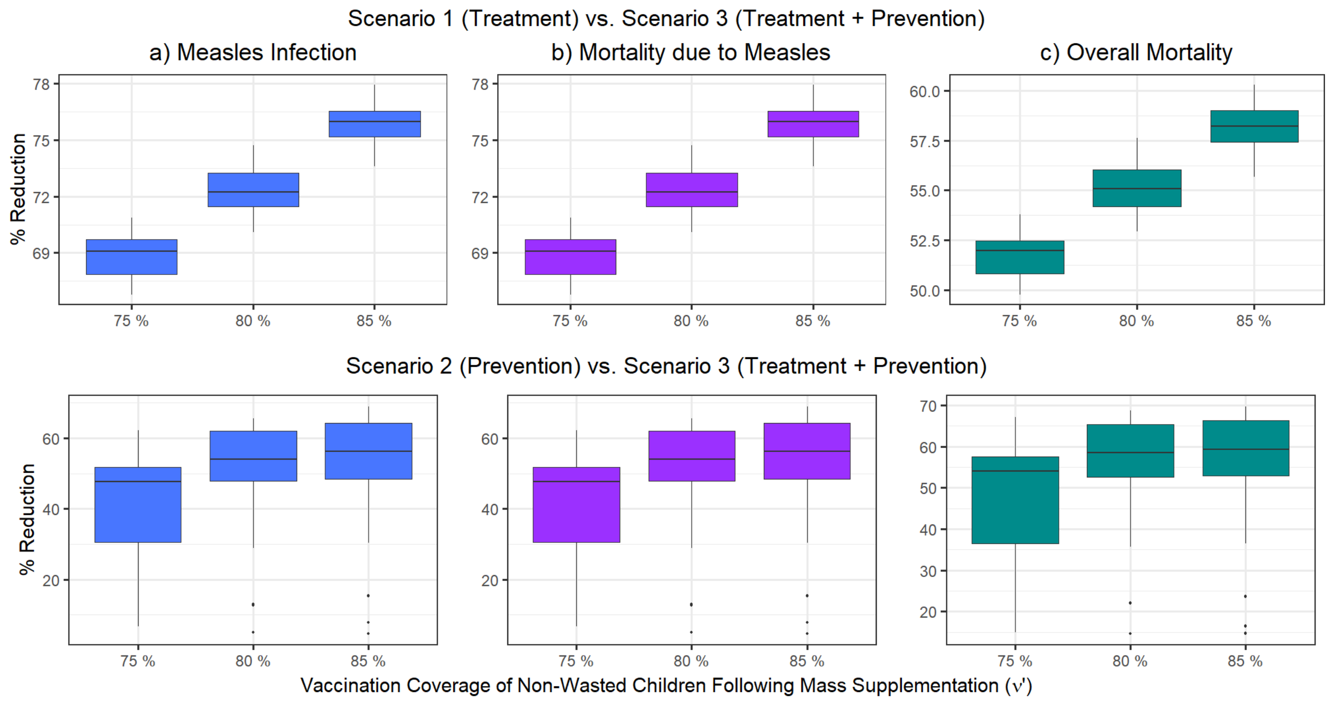


**Figure S 6.** Comparison of percentage reduction in measles infection and mortality due to measles across three scenarios. The variation in boxplots is produced by changing the treatment coverage of wasted children and vaccination coverage of children who received treatment for wasting.

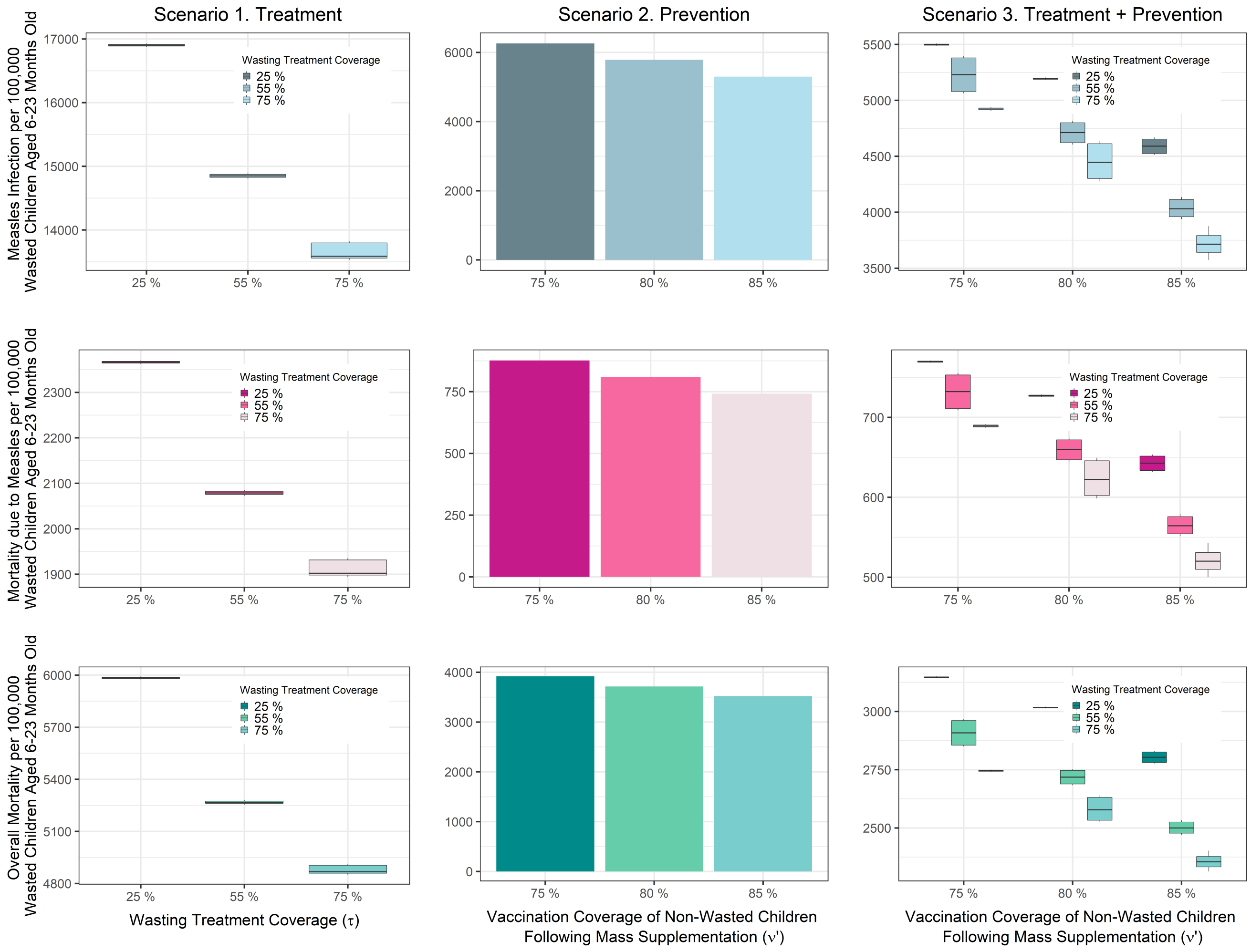


**Figure S 7.** Comparison of measles infection per 100,000 wasted children aged 6-23 months old, mortality due to measles per 100,000 wasted children aged 6-23 months old, and overall mortality per 100,000 wasted children aged 6-23 months old across three scenarios. Scenario 1 shows changes in outcomes by wasting treatment coverage of ($\tau$: 25%, 55% and 75%), and scenario 2 shows changes in outcomes by vaccination coverage of non-wasted children following mass supplementation ($\nu'$: 75%, 80%, 85%). Scenario 3 shows changes in outcomes by varying both $\tau$ and $\nu'$. The variation in boxplots is produced by changing the vaccination coverage of children who received treatment for wasting.

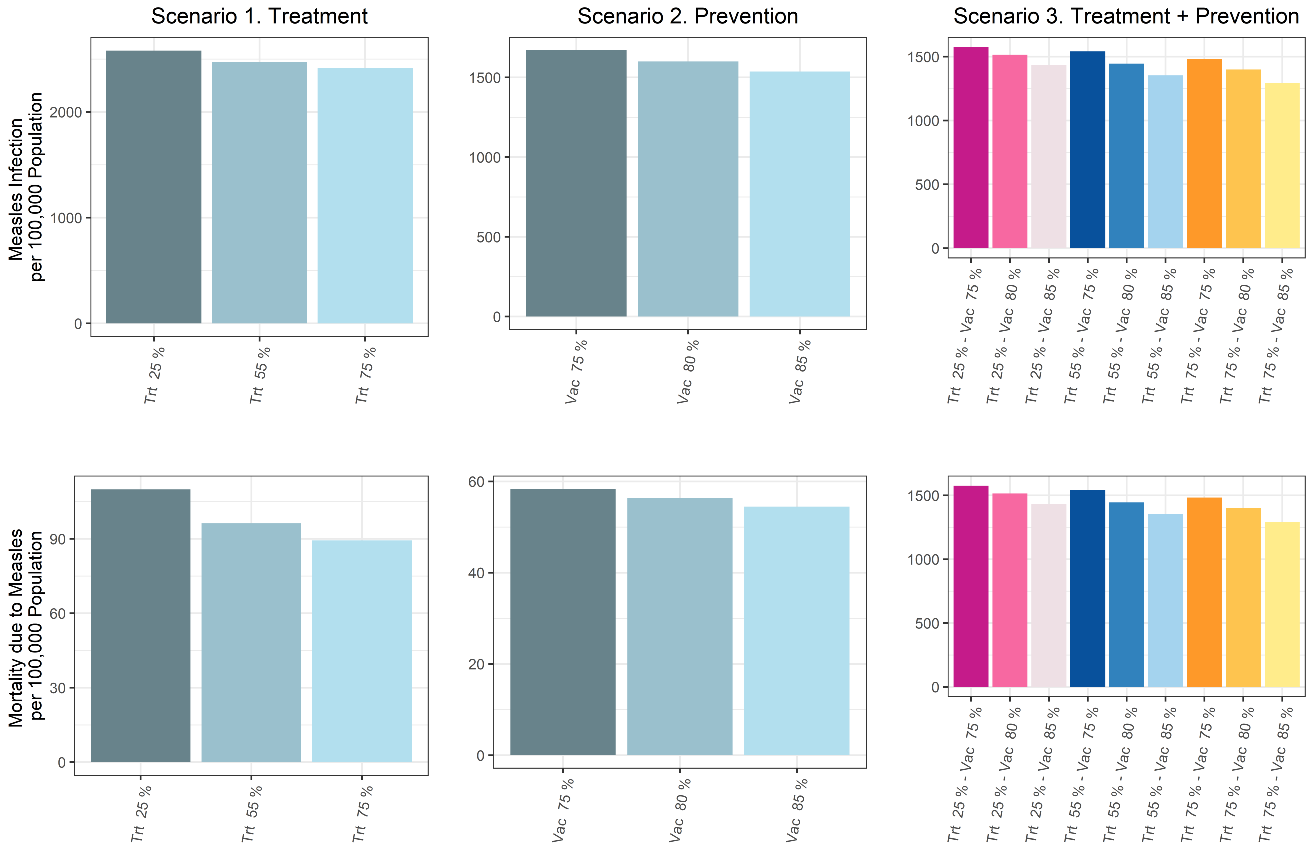


**Figure S 8.** Comparison of measles infection and mortality due to measles per 100,000 population across three scenarios. Vaccination coverage of children treated for wasting was fixed to $\nu t$: 75%. Scenario 1 shows changes in outcomes by wasting treatment coverage of ($\tau$: 25%, 55% and 75%), and scenario 2 shows changes in outcomes by vaccination coverage of non-wasted children following mass supplementation ($\nu'$: 75%, 80%, 85%). Scenario 3 shows changes in outcomes by varying both $\tau$ and $\nu'$.

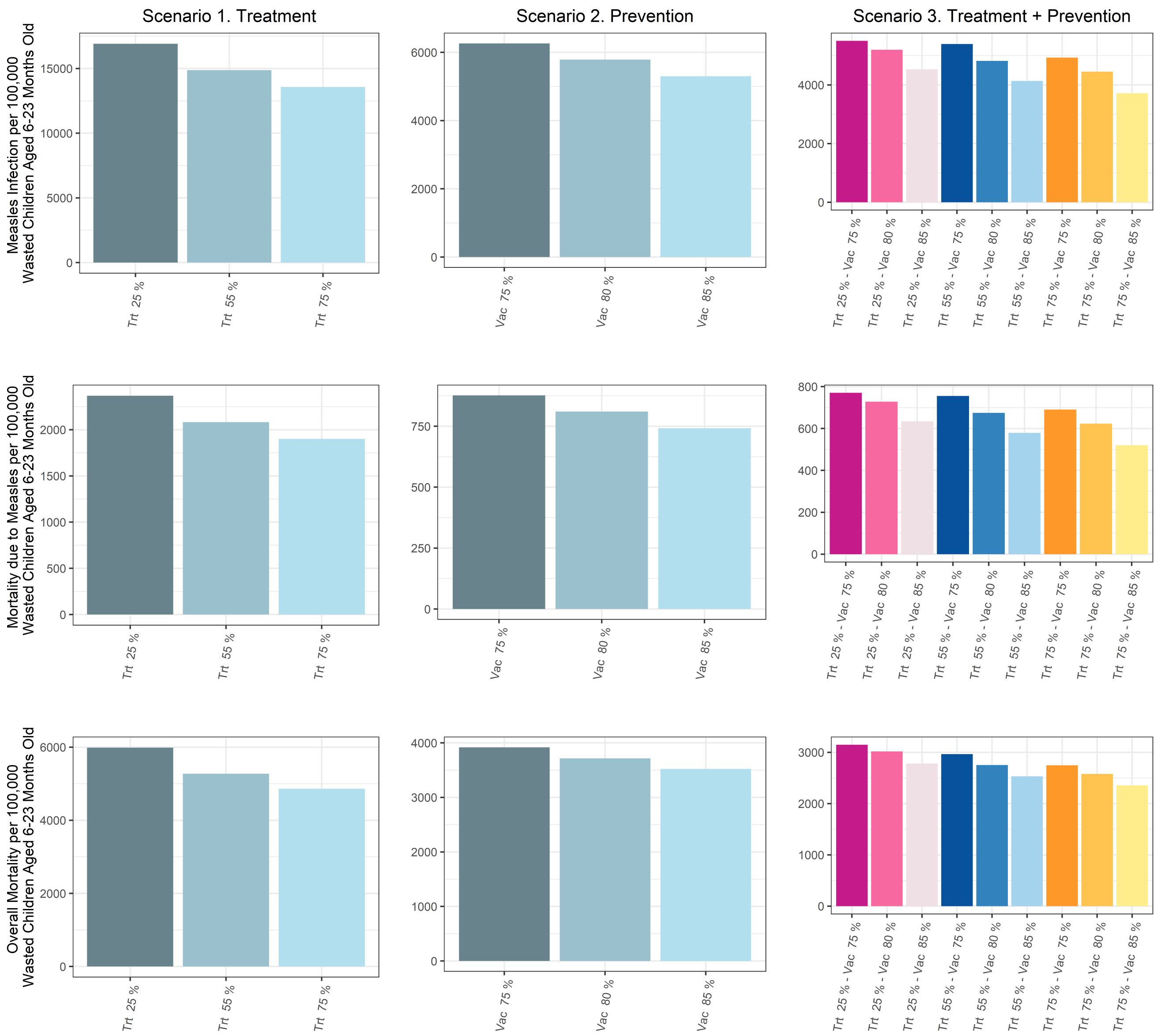


**Figure S 9.** Comparison of measles infection per 100,000 wasted children aged 6-23 months old, mortality due to measles per 100,000 wasted children aged 6-23 months old, and overall mortality per 100,000 wasted children aged 6-23 months old across three scenarios. Vaccination coverage of children treated for wasting was fixed to $\nu t$: 75%. Scenario 1 shows changes in outcomes by wasting treatment coverage of ($\tau$: 25%, 55% and 75%), and scenario 2 shows changes in outcomes by vaccination coverage of non-wasted children following mass supplementation ($\nu'$: 75%, 80%, 85%). Scenario 3 shows changes in outcomes by varying both $\tau$ and $\nu'$.

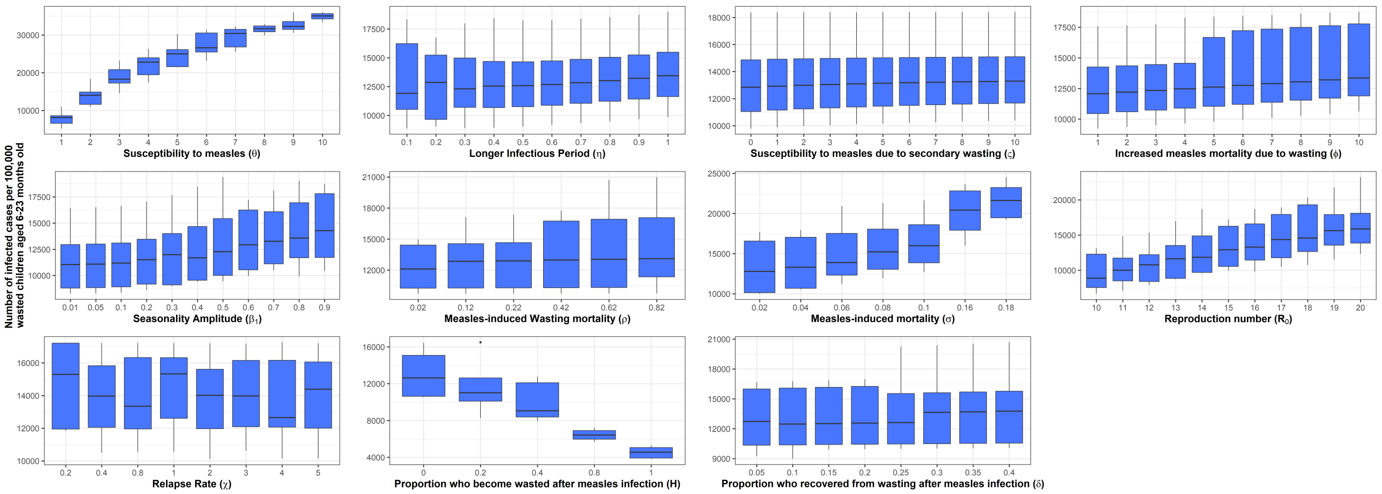


**Figure S 10.** Variation in measles infection per 100,000 wasted children aged 6-223 months old using wasting treatment (scenario 1), assuming the wasting treatment coverage ($\tau$) varies between 5% to 100% and vaccination coverage of treated children for wasting ($\nu t$) varies between 67.5% to 100%.

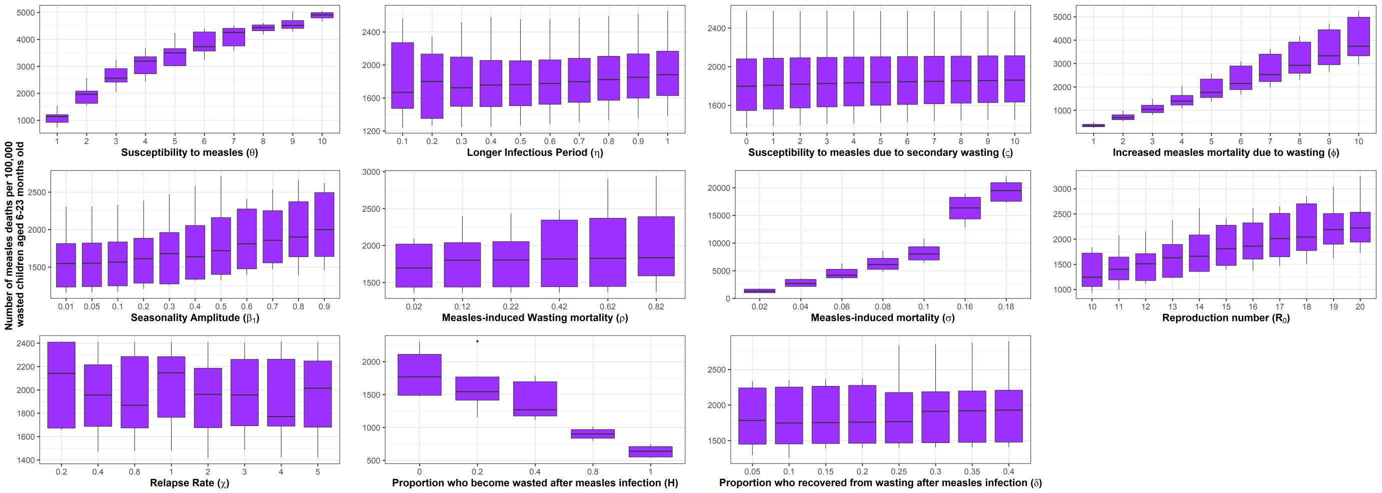


**Figure S 11.** Variation in mortality due to measles per 100,000 wasted children aged 6-223 months old using wasting treatment (scenario 1), assuming the wasting treatment coverage ($\tau$) varies between 5% to 100% and vaccination coverage of treated children for wasting ($\nu t$) varies between 67.5% to 100%.

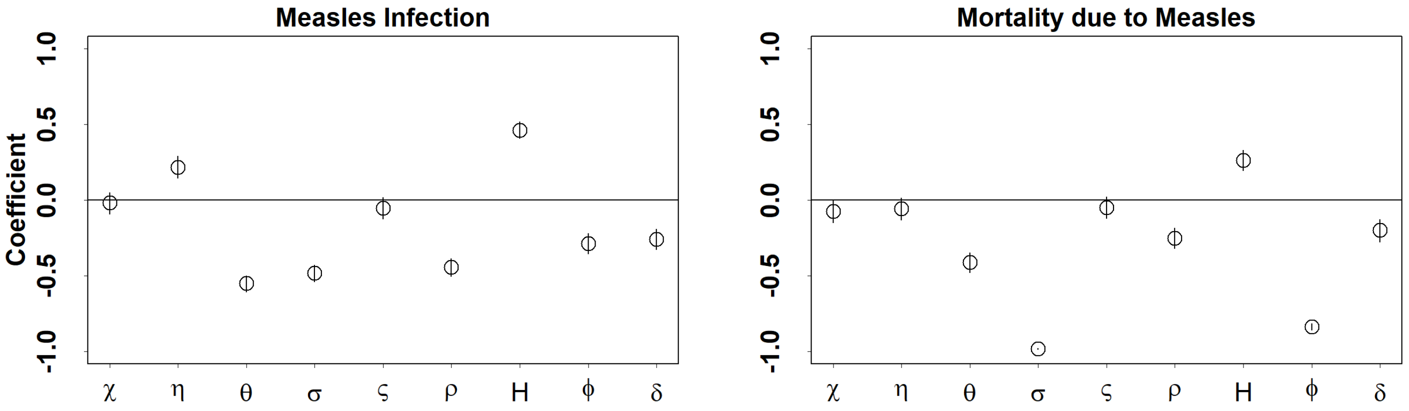


**Figure S 12.** Result of partial rank correlation coefficient of scenario 1 simulations using different model parameters. The further the coefficient of each parameter is from the horizontal line, the more sensitive the model outcome is to that parameter.
